# National Validation of a Dual-Outcome Risk Score for Trial of Labor After Cesarean: A Population-Based Analysis of 477,693 Deliveries

**DOI:** 10.64898/2026.04.07.26350334

**Authors:** Lauren Crabtree, Ciprian Paul Gheorghe

## Abstract

**Objective:** To externally validate, at the national level, a cumulative risk score for vaginal birth after cesarean (VBAC) success and neonatal morbidity derived from single-center data.

**Methods:** We conducted a population based cohort study of all trial of labor after cesarean (TOLAC) attempts among term, singleton deliveries recorded in the Centers for Disease Control and Prevention natality files, 2020 to 2024 (N=477,693). The cumulative risk score (range −1 to 7 points) incorporated body mass index (BMI) 30 or greater (+1), BMI 40 or greater (+1), induction of labor (IOL; +1), diabetes mellitus (+1), hypertensive disorder (+1), maternal age 40 years or older (+1), gestational age 41 weeks or greater (+1), and prior vaginal delivery (−1). VBAC success rates and neonatal intensive care unit (NICU) admission rates were evaluated across risk strata.

**Results:** The overall VBAC rate was 73.3% (350,340/477,693). The cumulative risk score demonstrated a monotonic relationship with VBAC success: score −1, 90.5%; score 0, 76.4%; score 1, 69.4%; score 2, 62.2%; score 3, 55%; and score 4 or higher, 44.8%. NICU admission rates increased concordantly from 43.8 to 111.1 per 1,000 across strata. Prior vaginal delivery was the strongest individual predictor (VBAC 86.4% vs 62.5%). VBAC rates and TOLAC volume were stable across 2020–2024.

**Conclusion:** The cumulative risk score derived from single-center data was externally validated in a national cohort of 477,693 TOLAC attempts. The monotonic dose-response relationship between risk score and both VBAC success and NICU admission was confirmed, supporting the use of this score for individualized TOLAC counseling.

## INTRODUCTION

Trial of labor after cesarean (TOLAC) represents a key decision point in obstetric care with implications at both the individual and population level. Successful vaginal birth after cesarean (VBAC) is associated with lower morbidity, whereas failed TOLAC is associated with higher risk than elective repeat cesarean delivery.^1–3^ In the United States, cesarean delivery now accounts for more than one in three births, creating a growing population of patients for whom subsequent delivery planning centers on the decision to pursue TOLAC.^4^ Across this group, pregnancy and delivery complications contribute billions of dollars in annual health care and societal costs, underscoring the importance of reducing preventable morbidity where safe alternatives to surgery exist.^5–7^ From a public health perspective, optimizing VBAC among appropriate candidates is a mechanism not only for improving maternal outcomes but also for mitigating the downstream costs of repeat operative delivery at scale.

Despite guideline support for TOLAC, access in the United States remains limited and geographically uneven.^8–10^ Hospitals offering TOLAC are clustered in a minority of US counties, with large regions of the country lacking local access, constraining the potential population level benefits of TOLAC and contributing to marked variation in VBAC rates that is unlikely to be explained by patient risk alone.^9–10^ There is therefore a need for tools that use routinely available clinical variables to provide clear estimates of both VBAC success and neonatal morbidity that are feasible for routine use in busy clinical settings to support patient centered counseling and informed decisions about repeat cesarean delivery.

Existing TOLAC prediction tools have important limitations that constrain their usefulness for system level implementation. Most population-based VBAC calculators were developed more than two decades ago in cohorts that predated the substantial increases in maternal age, obesity, and medical comorbidities that characterize contemporary obstetric populations.^11–14^ In this current era, these models may misclassify risk for a large share of TOLAC candidates, particularly those with more complex health profiles. Despite widespread use, existing VBAC calculators provide broad probability ranges limit both the precision of individual counseling and their clinical applicability, and they offer limited leverage for health systems seeking to standardize counseling and expand access to TOLAC at a population level.

In an institutional cohort of 1,418 consecutive TOLAC cases, a simple cumulative risk score was developed using seven routinely available clinical characteristics, with prior vaginal delivery treated as a protective factor.^15^ Across increasing score categories, VBAC success declined and neonatal intensive care unit (NICU) admission rose, indicating that the score stratified both successful TOLAC and neonatal morbidity using information available in routine clinical records.^15^ However, its single center derivation, modest sample size, and lack of external validation limited its suitability as a national dual outcome tool to guide practice and policy.

In this context, we sought to determine whether a simple, record-based TOLAC risk score retains its performance when applied across contemporary US practice. Using a nationally representative sample of term singleton TOLAC deliveries from 2020 through 2024, we sought to externally validate this cumulative risk score and determine whether the score can reliably stratify TOLAC risk in contemporary US practice and serve as a practical tool to support safe expansion of TOLAC at the population level.

## METHODS

### Study Design and Data Source

We performed a retrospective, population-based cohort study of all patients who underwent TOLAC among term singleton deliveries in the United States from 2020 through 2024, as captured in the Centers for Disease Control and Prevention (CDC) National Vital Statistics System (NVSS) natality files. Institutional review board approval was not required, as the dataset is publicly available and fully deidentified.

### Study Population

TOLAC was identified using two criteria applied in combination: the presence of at least one prior cesarean delivery (recorded in the risk factor checkbox on the birth certificate) and evidence of a trial of labor, defined as either a vaginal delivery after prior cesarean (successful VBAC) or a cesarean delivery with the trial of labor field marked as attempted (failed TOLAC). Patients who delivered by repeat cesarean without a documented trial of labor were classified as elective repeat cesarean and excluded. The cohort was restricted to term (37–42 weeks) singleton deliveries; the final analytic sample included 477,693 TOLAC attempts (N= 477,693).

### Cumulative Risk Score

The cumulative risk score was constructed identically to the index study.^15^ Points were assigned as follows: pre-pregnancy body mass index (BMI) 30 or greater (+1 point), BMI 40 or greater (+1 additional point), induction of labor (IOL; +1), diabetes mellitus (pregestational or gestational; +1), hypertensive disorder (chronic hypertension, gestational hypertension, or eclampsia; +1), maternal age 40 years or older (+1), and gestational age 41 weeks or greater (+1). Prior vaginal delivery was assigned −1 point, yielding a total score range of −1 to 7. Prior vaginal delivery was inferred from the difference between live birth order and number of prior cesarean deliveries; patients for whom this difference exceeded zero were classified as having had a prior vaginal delivery.

### Outcomes

The primary outcome was successful VBAC, defined as vaginal delivery (spontaneous, vacuum-assisted, or forceps-assisted) after a prior cesarean. The secondary outcome was NICU admission, as recorded on the birth certificate. In contrast to the index study, uterine rupture could not be assessed because it is not captured in the natality files; in the single center cohort, uterine rupture was included as a secondary outcome but was not reliably predicted by antepartum or intrapartum characteristics.^15^

### Statistical Analysis

VBAC success rates and NICU admission rates (per 1,000 deliveries) were calculated at each risk score level with Wilson 95% confidence intervals. Associations between individual risk factors and VBAC success were expressed as absolute differences in VBAC rates. The crude association between IOL and failed TOLAC was estimated as an odds ratio (OR) with 95% confidence intervals (CI). Logistic regression was used to evaluate predictors of VBAC success, and model discrimination was assessed by the area under the receiver operating characteristic curve (AUC). Temporal trends in VBAC rates and TOLAC volume were examined by year. Analyses were conducted in Python (version 3.10). Anthropic Claude (Anthropic) was used to assist with generation of Python code for data extraction and statistical analyses; all code output was reviewed, verified, and edited by the authors.

## RESULTS

### Cohort Characteristics

Among 18,209,300 deliveries recorded from 2020 to 2024, 477,693 met TOLAC inclusion criteria and comprised the analytic cohort (N= 477,693). Mean maternal age was 31.3 years (SD 5.1), mean pre-pregnancy body mass index was 28.1 (SD 6.7), and mean gestational age at delivery was 39.1 weeks (SD 1.2). Labor was induced in 29.6% of cases. Diabetes mellitus was present in 9.7% of patients, hypertensive disorders in 9.8%, and prior vaginal delivery in 45.4%. The overall VBAC rate was 73.3% (Table 1).

**Table 1.** *Cohort* Characteristics. Data are presented as N (%) or mean (SD).

| Characteristic | N (%) or Value |
| --- | --- |
| TOLAC cohort, N | 477,693 |
| VBAC success, N (%) | 350,340 (73.3) |
| Failed TOLAC, N (%) | 127,353 (26.7) |
| Maternal age, y, mean (SD) | 31.3 (5.1) |
| Pre-pregnancy BMI, kg/m<sup>2</sup>, mean (SD) | 28.1 (6.7) |
| Gestational age at delivery, wk, mean (SD) | 39.1 (1.2) |
| **Race and Ethnicity** |  |
| Non-Hispanic White | 222,027 (46.5) |
| Hispanic | 123,976 (26.0) |
| Non-Hispanic Black | 80,333 (16.8) |
| Non-Hispanic Asian | 30,320 (6.3) |
| Non-Hispanic Multiracial | 11,036 (2.3) |
| Non-Hispanic AIAN | 3,128 (0.7) |
| Non-Hispanic NHOPI | 2,021 (0.4) |
| **Number of Prior Cesarean Deliveries** |  |
| 1 | 426,904 (89.4) |
| 2 | 42,464 (8.9) |
| ≥3 | 7,611 (1.6) |
| **Risk Factors** |  |
| Induction of labor | 141,162 (29.6) |
| Diabetes mellitus | 46,456 (9.7) |
| Pregestational diabetes | 5,703 (1.2) |
| Gestational diabetes | 40,753 (8.5) |
| Hypertensive disorder | 46,956 (9.8) |
| Chronic hypertension | 12,668 (2.7) |
| Gestational hypertension | 33,682 (7.0) |
| Eclampsia | 773 (0.2) |
| Prior vaginal delivery | 216,863 (45.4) |
| Maternal age $\geq 40$ y | 24,755 (5.2) |
| BMI $\geq 30$ kg/m<sup>2</sup> | 152,706 (32.0) |
| BMI $\geq 40$ kg/m<sup>2</sup> | 27,471 (5.8) |
| GA $\geq 41$ wk | 56,847 (11.9) |
| NICU admission rate, per 1,000 | 58.4 |

### **R**isk Score and VBAC Success

VBAC success decreased steadily with increasing cumulative risk score across the full score range (Table 2). VBAC rates declined from 90.5% at score −1 (n=76,910) to 44.8% at score 4 or higher (n=7,644). Intermediate scores showed graded decreases in VBAC success between these extremes. The overall pattern and magnitude of risk across groups were consistent with those observed in the single-center derivation cohort, supporting the external validity of the risk score.^15^

**Table 2.**
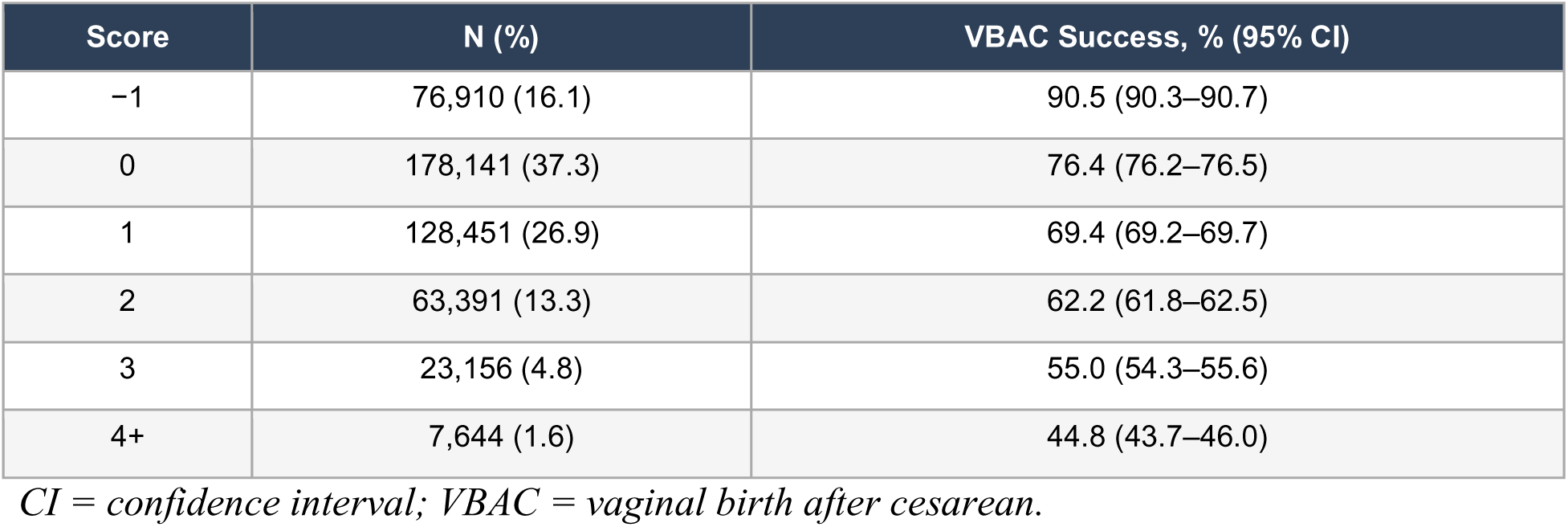
VBAC Success by Cumulative Risk Score. VBAC success rates with Wilson 95% confidence intervals.

### Neonatal Outcomes by Risk Score

NICU admission rates increased with higher cumulative risk scores (Table 3). Rates ranged from 43.8 per 1,000 at score −1 to 111.1 per 1,000 at score 4 or higher. Thus, higher risk scores in the national cohort were associated with both lower VBAC success and greater neonatal morbidity, consistent with the dual outcome framework underlying the risk score. The national NICU rates were generally lower in absolute terms than the institutional rates (43.8–111.1 vs 31.7–200.0 per 1,000), likely reflecting differences in NICU admission thresholds across the heterogeneous facilities represented in the national data.^15^

**Table 3.** NICU Admission by Cumulative Risk Score. NICU admission rates per 1,000 deliveries.

| Score | N (%) | NICU Admission, per 1,000 |
| --- | --- | --- |
| -1 | 76,910 (16.1) | 43.8 |
| 0 | 178,141 (37.3) | 51.2 |
| 1 | 128,451 (26.9) | 61.1 |
| 2 | 63,391 (13.3) | 73.0 |
| 3 | 23,156 (4.8) | 90.6 |
| 4+ | 7,644 (1.6) | 111.1 |

*NICU = neonatal intensive care unit.*

### Individual Risk Factor Associations

Prior vaginal delivery was the strongest individual predictor of VBAC success (86.4% vs 62.5%; Table 4). Among risk-increasing factors, hypertensive disorders were associated with the largest absolute reduction in VBAC rates (63.9% vs 74.4%), followed by BMI 40 or greater (58.6% vs 74.2%), and BMI 30 or greater (66.8% vs 76.4%). Diabetes mellitus (67.4% vs 74.0%) and induction of labor (70.8% vs 74.4%) were also associated with lower VBAC rates. Maternal age 40 years or older (70.7% vs 73.5%) and gestational age 41 weeks or greater (72.5% vs 73.4%) were each associated with modest absolute differences in VBAC success (Table 4). The rank order of risk factor effects was similar to that observed in the single-center index study.^15^

**Table 4.**
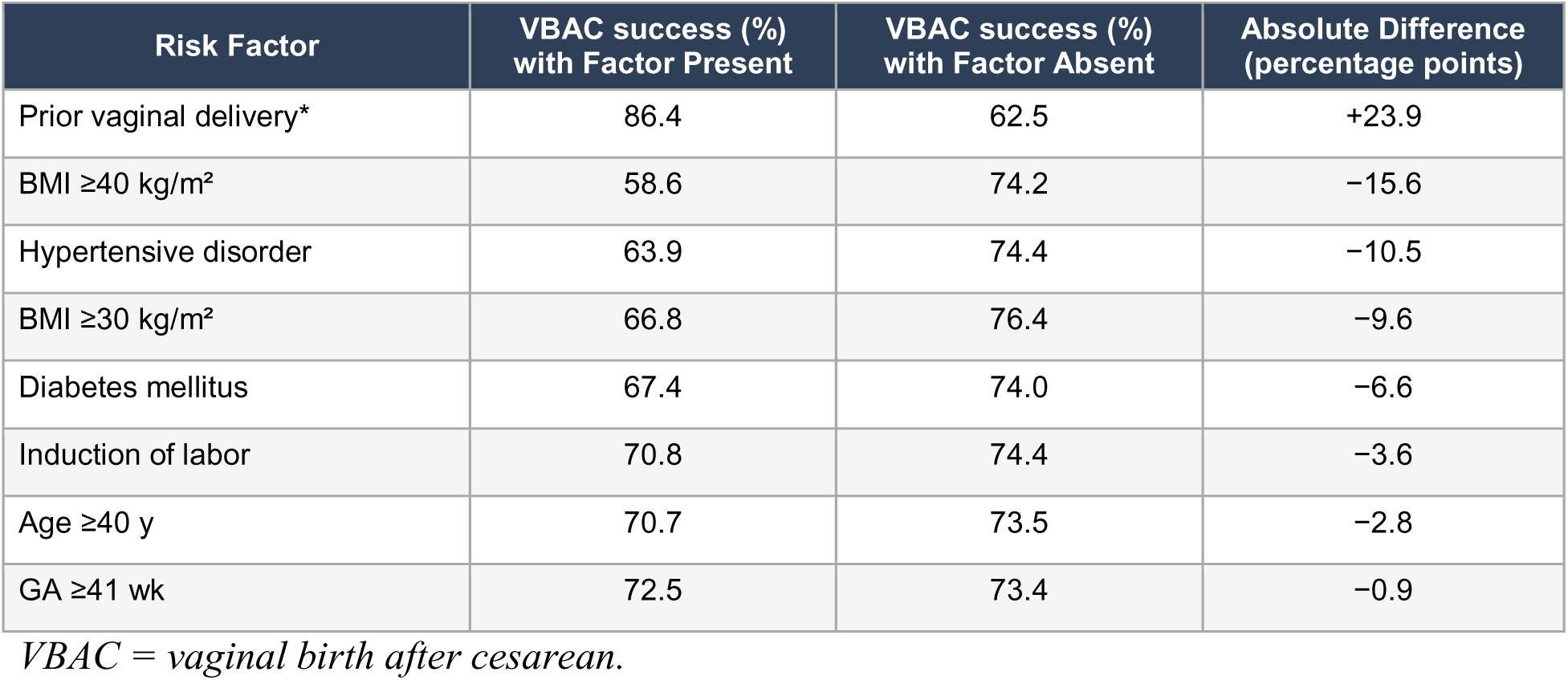
VBAC Success by Individual Risk Factor.

### Model Discrimination

Logistic regression on a 10% subsample (n=46,598) yielded an AUC of 0.574. This value was lower than the institutional AUC of 0.70, consistent with expected attenuation when a model derived from a clinically detailed single-center dataset is applied to administrative data lacking variables such as chorioamnionitis, indication for prior cesarean delivery, and Bishop score.^15^

### Temporal Trends

TOLAC volume increased from 89,615 in 2020 to 101,551 in 2024, a 13% increase over the study period. The VBAC rate was stable across all years (range 73.1% to 73.7%). NICU admission rates remained between 56.5 and 59.8 per 1,000.

## DISCUSSION

In this national analysis of 477,693 TOLAC attempts from 2020 through 2024, we externally validated a simple cumulative risk score, originally developed in a single tertiary center cohort, that stratifies both VBAC success and NICU admission across the full range of routinely collected ante/intrapartum factors.^15^ VBAC success decreased and NICU admission rates increased with higher risk scores, demonstrating clear separation of outcomes across score levels. Taken together, these findings suggest that a point-based, dual outcome score is a pragmatic tool for risk stratification and shared decision making and may help cautiously expand access to TOLAC among appropriate candidates in settings where labor after cesarean remains underused.

Prior vaginal delivery remained the strongest predictor of VBAC success at the population level. Patients with a prior vaginal delivery achieved VBAC in 86.4% of cases compared with 62.5% among those without, a 23.9–percentage point difference that exceeded the effect of any risk increasing factor. This supports weighting prior vaginal delivery as a protective (−1 point) element in the score and highlights patients with a prior vaginal birth as especially favorable candidates for TOLAC in routine counseling.

Among the risk increasing components of the score, hypertensive disorders and severe obesity (BMI 40 or greater) were associated with the largest absolute reductions in VBAC rates, followed by BMI 30 or greater, diabetes mellitus, induction of labor, maternal age 40 years or older, and gestational age 41 weeks or greater. The rank order of these effects paralleled the single center index study, reinforcing the construct validity of the point assignments and suggesting that the cumulative score captures a consistent gradient of TOLAC prognosis across diverse practice settings.^15^

Although this gradient was preserved, overall model discrimination was more modest in the national cohort than in the index study.^15^ In the national analysis, the AUC for VBAC prediction was 0.57 compared with 0.70 in the index study, a pattern consistent with applying a clinically detailed model to administrative data that lack key predictors and span heterogeneous practice environments.^15^ The strength of this cumulative score is that the same simple point system predicts both VBAC success and NICU admission risk, supporting its use for clinical counseling, particularly in settings where more granular clinical data (e.g., chorioamnionitis, indication for prior cesarean delivery, and Bishop score) are available, as in the index study.^15^

By applying the identical cumulative score to the complete United States natality files over five years, this analysis extends the original single center findings to a national TOLAC population and resolves the sparse cell counts that previously limited interpretation at the highest risk scores.^15^ The reproducible pattern of lower VBAC success and higher NICU admission at higher score levels suggests that the score generalizes to contemporary practice rather than representing a single institution phenomenon. These results are constrained by the limitations of birth certificate data, including the absence of indication and number of prior cesarean deliveries as clinically interpretable variables, lack of chorioamnionitis, Bishop score, and intrapartum management details, and the inability to ascertain uterine rupture, which was both clinically important and unpredictable in the institutional analysis. In addition, variation in NICU admission thresholds across hospitals likely contributes to the smaller differences in NICU admission rates observed nationally compared with the single□center estimates. The lower VBAC rates at intermediate scores (0 and 1) may reflect differences in which patients are offered TOLAC and how labor is managed at the tertiary center versus in broader United States practice.

In a setting where access to TOLAC remains uneven and many eligible patients never have the opportunity to attempt labor after cesarean, our findings support use of this simple cumulative score as a standard counseling tool.^8–10^ Using only routinely collected ante/intrapartum variables, this single score provides dual outcome estimates of VBAC success and NICU admission risk that are feasible to apply in busy clinical environments. Wider adoption of this framework could help expand access to TOLAC by identifying patients with favorable profiles while still recognizing those for whom elective repeat cesarean may be more appropriate. At the same time, any use of this score must remain anchored in the reality that TOLAC carries an inherent risk of uterine rupture and other acute complications, and should be offered only in settings with the resources to respond promptly to obstetric emergencies. Integrating this score into electronic health record calculators and standardized counseling workflows would offer a unified, dual□outcome alternative to existing VBAC tools and support more transparent, patient-centered discussions about risks and benefits in routine United States practice.

**Figure 1.**
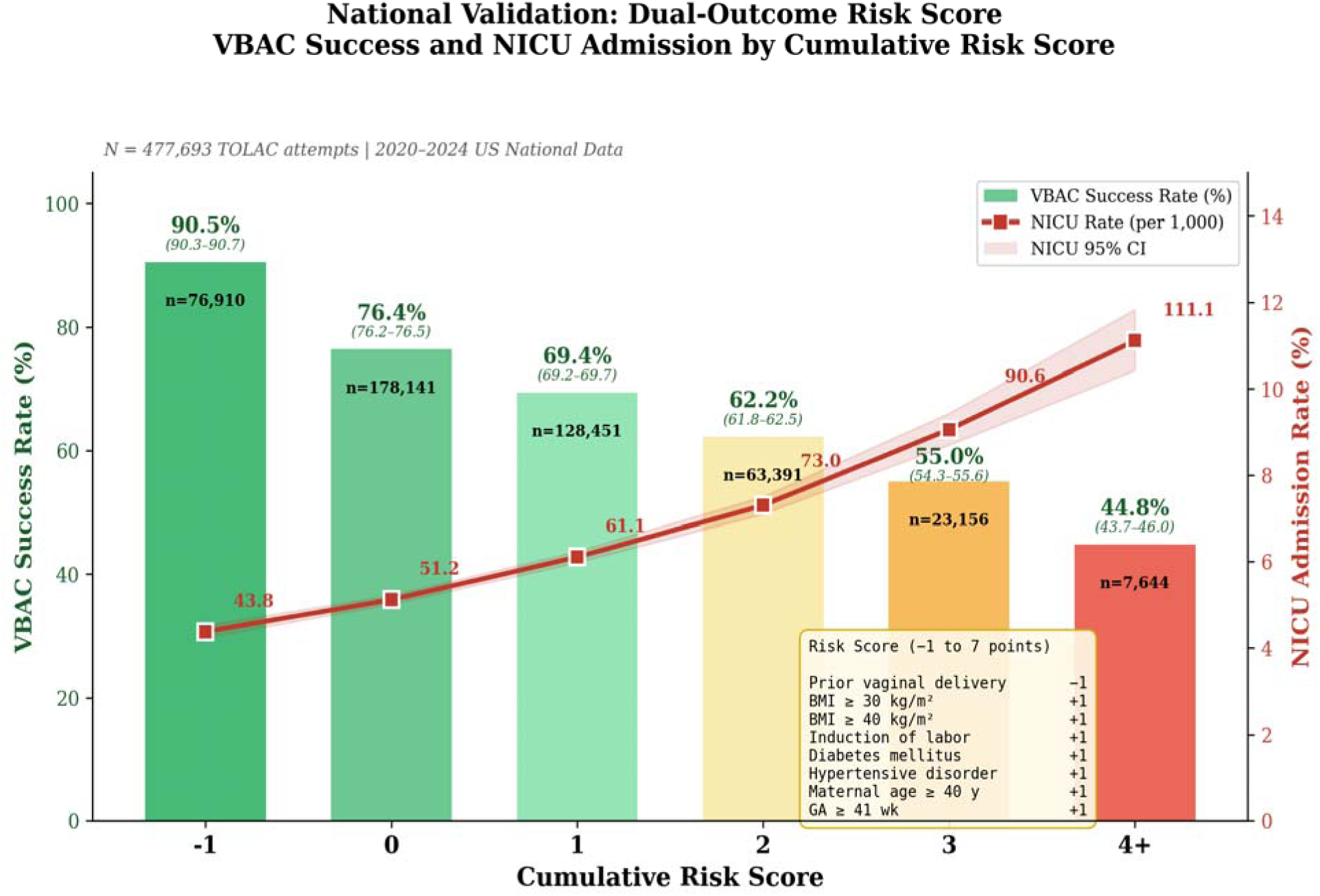
Dual-outcome relationship: VBAC success rate (bars, left axis) and NICU admission rate per 1,000 (line, right axis) by cumulative risk score in the national cohort. Both outcomes tracked concordantly across risk strata.

## Data Availability

All data produced in the present study are available upon reasonable request to the authors

## REFERENCES

1. American College of Obstetricians and Gynecologists. ACOG Practice Bulletin No. 205: Vaginal birth after cesarean delivery. Obstet Gynecol. 2019;133:e110–e127.

2. Landon MB, Hauth JC, Leveno KJ, et al. Maternal and perinatal outcomes associated with a trial of labor after prior cesarean delivery. N Engl J Med. 2004;351:2581–2589.

3. Lydon-Rochelle M, Holt VL, Easterling TR, Martin DP. Risk of uterine rupture during labor among women with a prior cesarean delivery. N Engl J Med. 2001;345:3–8.

4. Martin JA, Hamilton BE, Osterman MJK. Births in the United States, 2022. NCHS Data Brief. 2023;(477):1–8.

5. O’Neil SS, Platt I, Vohra D, et al. Societal cost of nine selected maternal morbidities in the United States. PLoS One. 2022;17(10):e0275656. Published 2022 Oct 26. doi:10.1371/journal.pone.0275656

6. Moran PS, Wuytack F, Turner M, et al. Economic burden of maternal morbidity - A systematic review of cost-of-illness studies. PLoS One. 2020;15(1):e0227377. Published 2020 Jan 16. doi:10.1371/journal.pone.0227377

7. Deshmukh U, Denoble AE, Son M. Trial of labor after cesarean, vaginal birth after cesarean, and the risk of uterine rupture: an expert review. Am J Obstet Gynecol. 2024;230(3S):S783–S803. doi:10.1016/j.ajog.2022.10.030

8. Chehab RF, Ferrara A, Grobman WA, et al. Racial, Ethnic, and Geographic Differences in Vaginal Birth After Cesarean Delivery in the US, 2011-2021. JAMA Netw Open. 2024;7(5):e2412100. doi:10.1001/jamanetworkopen.2024.12100

9. Ranchoff BL, Geissler KH, Goff SL, Bertone-Johnson ER, Paterno MT, Attanasio LB. Trends in Labor After Cesarean Delivery Access in the US. JAMA Netw Open. 2025;8(8):e2526224. doi:10.1001/jamanetworkopen.2025.26224

10. Robinson S, Royer H, Silver D. Geographic Variation in Cesarean Sections in the United States: Trends, Correlates, and Other Interesting Facts. J Labor Econ. 2024;42(Suppl 1):S219–S259. doi:10.1086/728804

11. Grobman WA, Lai Y, Landon MB, et al. Development of a nomogram for prediction of vaginal birth after cesarean delivery. Obstet Gynecol 2007;109:806–812.

12. Grobman WA, Lai Y, Landon MB, et al. Can a prediction model for vaginal birth after cesarean also predict the probability of morbidity related to a trial of labor? Am J Obstet Gynecol 2009;200:56.e1–56.e6.

13. Ashwal E, Wertheimer A, Aviram A, Wiznitzer A, Yogev Y, Hiersch L. Prediction of successful trial of labor after cesarean - the benefit of prior vaginal delivery. J Matern Fetal Neonatal Med. 2016;29(16):2665–2670. doi:10.3109/14767058.2015.1099156

14. Guise JM, Eden K, Emeis C, et al. Vaginal birth after cesarean: new insights. Evid Rep Technol Assess (Full Rep) 2010;191:1–397.

15. Crabtree L, Gheorghe CP. A novel dual-outcome risk calculator for trial of labor after cesarean. medRxiv. Preprint. Posted online March 20, 2026. doi: 10.64898/2026.03.18.26348725

